# Space-Earth connection: How space weather fluctuations impact epigenetic aging in an elderly men cohort from Massachusetts, USA

**DOI:** 10.1101/2025.11.11.25340039

**Authors:** Ting Zhai, Carolina L Zilli Vieira, Pantel Vokonas, Andrea A. Baccarelli, Zachary D. Nagel, Joel Schwartz, Petros Koutrakis

## Abstract

**Background:** In a previous study, we reported associations between space weather [galactic cosmic rays (GCRs)] and solar and geomagnetic activities (SGAs)] with shorter telomere length in a cohort of elderly men in Massachusetts. Here, we investigated the impact of space weather on epigenetic aging in the same cohort.

**Methods:** We analyzed 1,487 blood DNA methylation measures from 771 older men in the Normative Aging Study (1999–2013). Daily space weather indicators were obtained from NASA including sunspot number (SSN) and interplanetary magnetic field as solar activity parameters, Kp-index as a geomagnetic parameter, and neutron monitors and modeled cosmic ray-induced ionization as measures of GCRs. The 30-day moving average of each parameter was prespecified as the exposure window. Four epigenetic age acceleration metrics, including HorvathAgeAccel, HannumAgeAccel, PhenoAgeAccel, and GrimAgeAccel, were derived, and exploratory epigenome-wide association study (EWAS) and pathway enrichment analyses were conducted.

**Results:** GCRs were associated with accelerated epigenetic aging, whereas SGAs were associated with slower aging. Each interquartile range increase in SSN corresponded to a 0.61-year lower HorvathAgeAccel and 0.50-year lower PhenoAgeAccel, while higher neutron counts were associated with 0.32-year greater HorvathAgeAccel and 0.29-year greater HannumAgeAccel. EWAS identified hundreds of CpGs associated with GCRs (predominantly lower methylation) and thousands with SGAs (predominantly higher methylation), enriched in genome maintenance pathways such as P53 signaling, DNA repair, and inflammatory response, consistent with astronaut studies showing activation of similar stress and repair pathways.

**Conclusion:** Short-term space weather fluctuations were associated with distinct epigenetic aging patterns in blood, suggesting that, as observed in astronauts, terrestrial populations may likewise show biological sensitivity to space weather variability.

## Introduction

The general population at ground level is continuously exposed to low-level environmental ionizing and non-ionizing radiation linked to space weather, including solar and geomagnetic activities (SGAs) and galactic cosmic rays (GCRs). Primary GCRs penetrate the Earth’s atmosphere and induce ionization through a nuclear-electromagnetic-muon cascade, resulting in secondary particles such as muons, neutrons, and cosmic ray-induced ionization (CRII). Long-term follow-up studies of the general population have reported associations between space-weather variability and multiple adverse aging-related health outcomes, including impaired lung function,^1^ cognitive disfunction,^2^ cardiovascular disease (CVD) morbidity and mortality.^3–6^ These findings highlight the potential public health burden of space-weather exposures in general population, but the biological mechanisms remain poorly understood.

In our prior work, we showed that GCR exposure was associated with telomere shortening, reflecting altered genome stability.^7^ Ionizing radiation exposure even at low levels can induce DNA damage and oxidative stress,^8^ thereby activating genome maintenance pathways such as DNA repair and telomere regulation. In this study, we hypothesize space weather may affect aging processes as reflected in DNA methylation (DNAm). DNAm represents a dynamic and environmentally responsive epigenomic marker, in contrast to genomic alterations that typically accumulate more gradually over the lifespan and are less responsive to short-term variation.^9^ Because DNAm regulates gene expression and cellular function, changes in methylation patterns can capture both early responses to exposures and longer-term reprogramming.^10^ Notably, measurable shifts in DNAm have been observed within days to months in relation to air pollution, smoking, and diet.^11–13^ Radiation-related DNAm alterations have also been observed in high-dose settings such as astronauts and radiotherapy patients.^14–20^

Although radiation dose rates at the ground level are far lower than in orbit, the background oscillations of space weather on ∼11-year solar cycles create a natural experiment where small but persistent electromagnetic radiation exposures may leave detectable signatures in the epigenome.^7^ Moreover, day-to-month fluctuations in space weather could plausibly induce short-term DNAm changes, which in turn may contribute to cumulative biological aging and aging-related disease risk. DNAm-based epigenetic clocks have emerged as robust biomarkers^21,22^ that capture different aspects of biological aging: first-generation clocks such as HorvathAge and HannumAge reflect chronological and intrinsic cellular aging, while newer models such as PhenoAge and GrimAge incorporate clinical or mortality-related information to better represent systemic physiological decline.^21,23–25^ The difference between DNAm-predicted and chronological age, known as epigenetic age acceleration (EAA), has widely been associated with elevated risk of age-related disease and mortality.^26–28^ Together, these tools provide an established framework to test whether space-weather variability influences biological aging in the general population.

In this study, we analyzed the association between short-term space-weather fluctuation with epigenetic aging and DNAm changes. We analyzed data from the Normative Aging Study (NAS), a longitudinal cohort of older adults with repeated DNAm measures, relating indicators of GCR and SGA to (i) EAA across established aging clocks as the primary outcome; and (ii) exploratory analyses of global and context-specific methylation entropy as well as differentially methylated CpG sites and regions.

## Methods

### Study population

NAS is a longitudinal cohort of older men established in 1963 by the U.S. Department of Veterans Affairs. NAS participants were initially free of chronic diseases at enrollment and have been followed prospectively with repeated health examinations. In this study we included 771 participants with 1487 visits with complete DNAm data between 1999 and 2013. The follow-up visits occurred approximately every 3 years. At each visit, demographic and clinical characteristics were collected through routine physical examinations (height and weight), laboratory tests (white blood cell count, percent neutrophils, percent lymphocytes), collection of medical history information (treatment with statin medication, treatment with hypertension, diagnosis of diabetes), and completion of questionnaires on smoking history, education level, and other factors that may affect health. Body mass index (BMI) was computed as the weight in kilograms divided by the square of the height in meters based on height and weight measurements at each visit. This study was approved by the institutional review boards of the Harvard T.H. School of Public Health and the Veterans Administration Boston Healthcare System, and all participants provided written informed consent.

### Exposure assessment

The space weather assessment has been described in our previous study.^7^ We used sunspot number (SSN) and interplanetary magnetic field (IMF) intensity as indicators of solar activity along with Kp-index as indicator of geomagnetic disturbances. These data were obtained from the NASA Goddard Space Flight Center and were converted from Coordinated Universal Time to Eastern Time, the time zone in Boston, MA. GCRs at the ground level were estimated by the sum of paired ions cm^−3^ sec^−1^ produced by primary and secondary GCRs in atmospheric reactions with the CRII model as previously described by Usoskin and Kovaltsov.^29^ Neutrons were estimated from the neutron monitors in the Bartol Research Institute, University of Delaware, USA.^30^ The moving averages for the space weather indicators were calculated by taking the average of the daily measures before the date of each DNAm measurement, including 7-day, 30-day, and 180-day moving averages.

### DNA methylation (DNAm)

DNAm was measured by the Infinium HumanMethylation450 BeadChip (Illumina, San Diego, CA) with genomic DNA extracted from whole blood buffy coat using standard procedures.^11^ The samples were randomized across plates and assigned through a two-stage age-stratified algorithm to minimize batch effects and maintain a similar age distribution across batches. Detailed quality control of samples and data has been previously described.^31,32^ Briefly, methylation data were corrected for dye bias, and probes below background fluorescence (detection p-value < 0.01), outliers, sex-chromosome probes, non-CpG probes, SNP-associated probes, cross-reactive probes, and non-unimodal probes were removed. After QC, 360,272 high-quality probes entered the epigenome-wide association study (EWAS) (dbGaP accession: phs000853.v2.p2). The methylation data were normalized using the beta-mixture quantile normalization (BMIQ) method implemented in the ChAMP package,^33,34^ and batch effects were estimated using the sva package with 8 surrogate variables optimized using num.sva to control the inflation factor of downstream EWAS.^35^ Probes were annotated with IlluminaHumanMethylation450kanno.ilmn12.hg19.

We quantified normalized methylation entropy (NME) both genome-wide and within different genomic contexts, including OpenSea, CpG islands, and their flanking regions (north and south shores and shelves). Specifically, for each CpG site, NME reflects the degree of disorder or uncertainty in methylation by considering the distribution of methylated and unmethylated states. ^16^ These site-level entropy values were averaged across CpGs and rescaled to a 0-1 scale, with higher values indicating more variable or less ordered methylation patterns.

### Epigenetic aging biomarkers

Epigenetic aging clocks were calculated using the online platform hosted by the Clock Foundation (https://dnamage.clockfoundation.org).^21^ We focused on four widely validated clocks: HorvathAge, HannumAge, PhenoAge, and GrimAge, due to their well-established ability to predict chronological age and age-related outcomes. Clock estimates were based on normalized beta values following QC. EAA was defined as the residual from regressing each clock estimate on chronological age, while adjusting for batch effect indicators and estimated blood cell type compositions (CD8+ T cells, CD4+ T cells, B cells, natural killer cells, monocytes, and granulocytes).^22^

### Statistical analysis

Linear mixed effect models (LME) were performed for phenotype level associations. A random intercept was included to account for repeated measures within participants. We specified three sequential covariate adjustment models: (1) adjusting for age, year, and methylation technical factors (cell type composition and batch effect indicators); (2) additionally adjusting for participant characteristics, including smoking status, pack-years, BMI, years of education, diabetes, hypertension medication, and statin; and (3) further adjusting for environmental variables (temperature, visibility, and season) in addition to model 2 covariates. Models were fit for each space weather indicator separately to avoid collinearity and to evaluate exposures independently. Each indicator was scaled by its interquartile range (IQR), and the effect estimates were reported as changes per IQR increase to facilitate cross-indicator comparison and enhance interpretability.

As a sensitivity analysis, we assessed the association of EAA metrics with alternative exposure windows of GCRs and SGAs (daily, 7-day, and 180-day moving averages) to evaluate robustness and biological plausibility of the prespecified 30-day moving average, which was selected based on biological rationale and prior studies of environmental influences on the epigenome.^11,36^ Daily, 7-, and 180-day results are reported in sensitivity analyses. Natural cubic spline models with 3 degrees of freedom were fit to explore potential non-linear relationships between each space weather indicator and EAA.

### Epigenome-wide association study (EWAS)

EWAS was performed using the limma package with empirical Bayes estimation. The response variable was the methylation level expressed as m-values (logit-transformed beta values). The predictor of interest was each space weather indicator, adjusting for the same full covariate sets described in the *Statistical analysis* section. To account for repeated measures, we estimated duplicate correlations using participant ID as the blocking factor and incorporated consensus correlations (duplicateCorrelation function) into the limma model. Since 360,272 tests were performed for each exposure, bacon correction followed by Benjamini-Hochberg (BH) false discovery rate (FDR) correction was applied, with significance defined at FDR < 0.05. EWAS inflation factors were all <1.15.

### Region and pathway analysis

Differentially methylated region (DMR) analysis was performed using the comb-p method,^37^ with CpG-level statistics from the EWAS as input, applying a seed p-value threshold of 0.001 and distance cutoff of 1,000 bp. Regions were considered significant if FDR < 0.05 and containing ≥ 3 contributing CpGs. We further defined a hotspot as any 1-kb window containing ≥ 1 significant comb-p DMR for ≥ 2 indicators. Hotspots were then ranked by number of indicators and minimum region FDR. For each hotspot x indicator, we plotted CpG-level moderated t-statistics and assigned direction by the sign of t. Overlap across indicators was computed using ≥ 1 bp intersection, with a reciprocal-overlap sensitivity.

Gene-set enrichment analysis was conducted using methylGSA,^38^ which accounts for CpG-to-gene mapping multiplicity and probe-count bias. Primary analyses used bacon- and BH-corrected CpG FDRs as input to methylGSA’s p-value based tests (mGLM and mRRA), testing Gene Ontology (GO: BP, CC, MF) categories and MSigDB Hallmark gene sets (human, version 7.1). We prespecified 14 genome integrity related Hallmark sets (DNA Repair, Reactive Oxygen Species, P53, Oxidative Phosphorylation, Peroxisome, mTORC1 Signaling, PI3K-AKT-mTOR Signaling, Unfolded Protein Response, Protein Secretion, Apoptosis, TNFA via NFKB, Interferon-γ Response, Interferon-α Response, Inflammatory Response). Because the EWAS showed pronounced directional asymmetry, we also performed secondary analyses splitting CpGs into higher versus lower methylation subsets and repeated enrichment to characterize directional asymmetry.

All analyses were conducted in R version 4.3.3.

## Results

In total, 771 participants contributed to 1487 visits with complete DNAm data during 1999 and 2013, with up to 4 total visits. The participants were all male, with a mean age of 72.64 (6.76) years at baseline (**Table 1**). Daily levels of space-weather indicators across the study period showed peaks consistent with solar cycles: SSN peaked around 2000-2002 and again in the early 2010s; IMF and Kp showed similar trends, whereas CRII and neutron counts were anti-phase (**Fig. 1a**). IQRs of each space weather indicator are summarized in **Table 2**.

**Figure 1.**
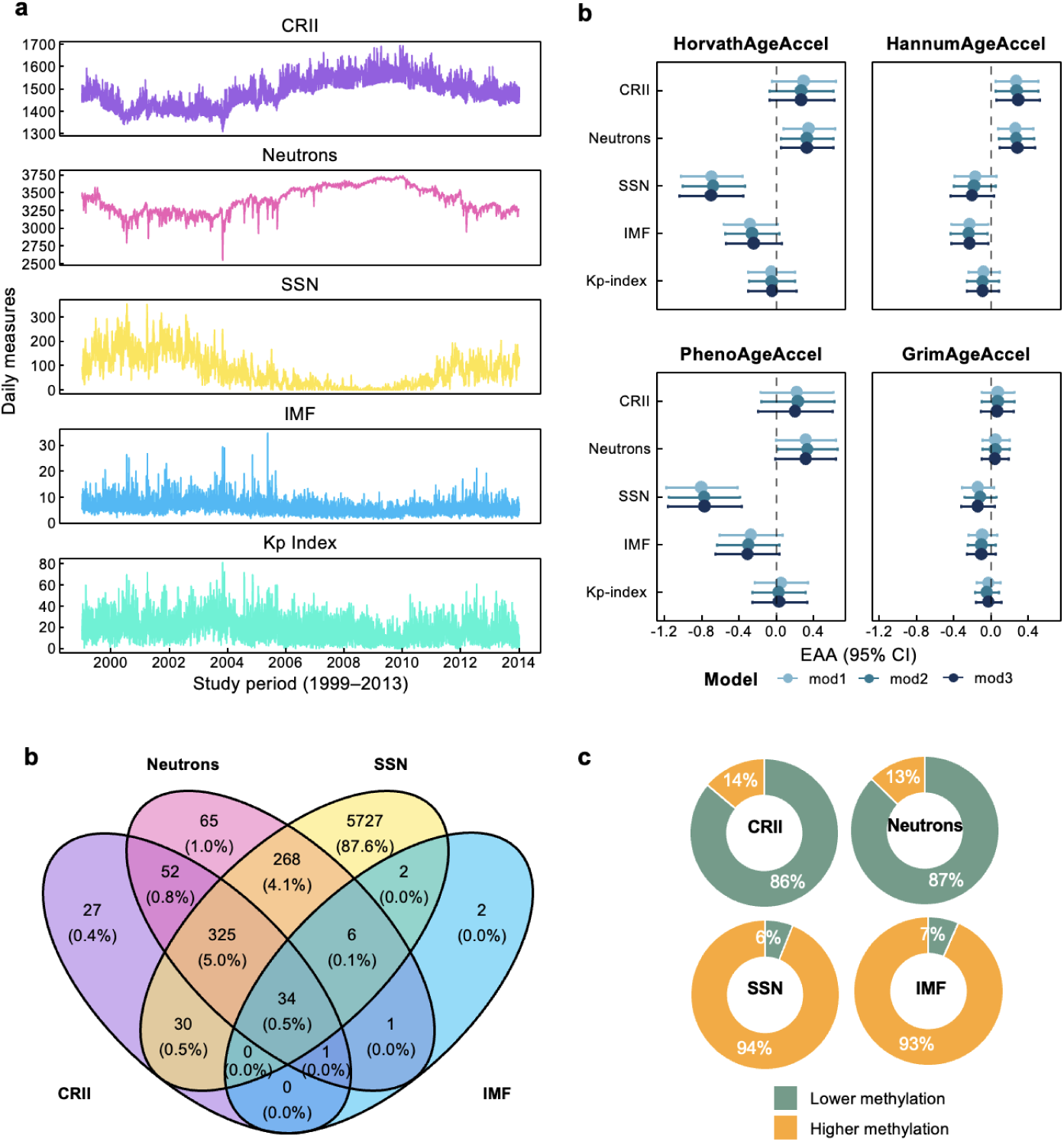
Space-weather exposures and epigenome-wide association study (EWAS) results. **(a)** Daily values of solar and geomagnetic activity indices, including sunspot number (SSN), interplanetary magnetic field (IMF) intensity, and Kp index, and ground-level proxies of galactic cosmic rays (GCRs), including cosmic ray-induced ionization (CRII) and neutron counts, measured during the Normative Aging Study (NAS) DNA methylation assessment period (1999-2013). **(b)** Associations between space-weather exposures (per interquartile range of 30-day moving average) and epigenetic age acceleration (EAA), including HorvathAgeAccel, HannumAgeAccel, PhenoAgeAccel, and GrimAgeAccel. Results are presented as β estimates (change of EAA in years) with 95% confidence intervals (CIs). Models were specified sequentially: mod1 adjusted for age, year, blood-cell composition, and batch factors; mod2 additionally adjusted for smoking status, pack-years, body mass index, education, diabetes, antihypertensive medication, and statin use; mod3 further adjusted for temperature, visibility, and season. **(c)** Numbers of differentially methylated CpG sites identified in EWAS (false discovery rate [FDR] < 0.05 with bacon correction and Benjamini-Hochberg adjustment), shown as unique or overlapping across CRII, neutrons, SSN, and IMF. **(d)** Proportions of CpGs showing lower methylation versus higher methylation by exposure type. EWAS were conducted separately for each exposure, scaled per interquartile range (IQR) of its 30-day moving average. Models were adjusted for age, year, blood-cell composition, batch factors, smoking status, pack-years, body mass index, education, diabetes, antihypertensive medication, statin use, temperature, visibility, and season. The Kp index did not yield significant CpGs at FDR < 0.05 and was excluded from downstream analyses.

**Table 1.** Baseline and longitudinal characteristics of participants.

| Variable | Baseline<br>(N = 771) | All visits<br>(N = 1,487) |
| --- | --- | --- |
| Age (years) | 72.64 (6.76) | 74.51 (6.97) |
| BMI (kg/m<sup>2</sup>) | 28.16 (4.07) | 28.01 (4.11) |
| Smoking status |  |  |
| Never | 236 (30.9) | 465 (31.4) |
| Current | 33 ( 4.3) | 63 ( 4.3) |
| Former | 495 (64.8) | 952 (64.3) |
| Packyears | 12 (35) | 12 (34) |
| Education years | 15.01 (2.95) | 15.13 (3.00) |
| Diabetes | 104 (13.5) | 223 (15.0) |
| Hypertension | 451 (58.5) | 945 (63.6) |
| Statin use | 270 (35.0) | 658 (44.3) |
| WBC (10<sup>9</sup>/L) | 6.31 (1.66) | 6.36 (1.70) |
| Neutrophils % | 62.25 (8.10) | 62.60 (8.16) |
| Lymphocytes % | 25.36 (7.30) | 24.87 (7.30) |

Values are mean (SD) for continuous variables or n (%) for categorical variables. Pack-years presented as median (IQR). Abbreviations: BMI = body mass index; WBC = white blood cell count.

**Table 2.** Interquartile ranges of space weather indicators by averaging window.

| Variable | Daily | 7-day | 30-day | 180-day |
| --- | --- | --- | --- | --- |
| CRII (ion pairs cm<sup>-3</sup> s<sup>-1</sup>) | 91.4 | 89.8 | 88.5 | 77.4 |
| Neutrons | 235.2 | 227.3 | 226.1 | 218.1 |
| SSN | 115.4 | 110.5 | 110.1 | 105.9 |
| IMF (nT) | 2.95 | 2.13 | 1.69 | 1.17 |
| Kp index | 16.4 | 9.04 | 7.19 | 3.46 |

Values represent interquartile ranges (IQRs) of daily, 7-day, 30-day, and 180-day moving averages prior to each DNA methylation assessment. Abbreviations: CRII = cosmic ray induced ionization; SSN = sunspot number; IMF = interplanetary magnetic field.

### Epigenetic aging clocks

How epigenetic aging and its acceleration fluctuated over the study period is illustrated in participants with at least three repeated samples (**Fig. S1**). We examined first whether fluctuations in space weather indicators were associated with accelerated epigenetic aging. We found clear and opposite patterns consistent with our prior findings in telomere length: SGAs were associated with lower EAA, while GCRs were associated with higher EAA. Each IQR increase in the 30-day moving average of SSN was associated with a 0.61-year lower HorvathAgeAccel (95% CI: −0.98, −0.23) and a 0.50-year lower PhenoAgeAccel (95% CI: −0.94, −0.07) in the fully adjusted models. Similarly, IMF was associated with lower HannumAgeAccel (−0.25 years per IQR, 95% CI: −0.45, −0.05) and borderline lower PhenoAgeAccel. In contrast, higher neutron intensity was associated with higher HorvathAgeAccel (0.32 years per IQR, 95% CI: 0.04, 0.60) and HannumAgeAccel (0.29 years per IQR, 95% CI: 0.10, 0.48), with borderline higher PhenoAgeAccel. Additionally, CRII increase was associated with higher HannumAgeAccel (0.31 years per IQR, 95% CI: 0.08, 0.54) (**Fig. 1b**). No clear associations were observed for GrimAgeAccel or Kp-index.

While effect sizes and significance varied across clocks, indicators within the same category (SGAs vs. GCRs) showed consistent directional trends, suggesting that each clock captures partly distinct but related aspects of biological aging influenced by these exposures. These opposite associations between SGAs and GCRs parallel our previous telomere findings, where SGAs predicted longer telomeres and GCRs shorter ones, consistent with their known anticorrelation in solar cycles.^7,39^ Sensitivity analyses showed that the 30-day moving average associations were largely linear as explored by the spline models (**Fig. S2**). Results were directionally consistent using daily, 7-day, and 180-day windows, although with attenuated magnitudes (**Fig. S3**).

### Methylation entropy patterns

We next summarized variability of methylation at global level and the context level using the normalized entropy across CpGs. SGAs were associated with greater variability (higher entropy) in CpG islands, whereas GCR proxies mainly reduced entropy in islands (**Fig. S4**). Per IQR increase in SSN (30-day moving average), island entropy increased by 0.13 percentage points (pp) (95% CI: 0.09, 0.18), suggesting more variable promoter-proximal methylation under higher SGA activity. CRII decreased island entropy by 0.04 pp (95% CI: −0.08, 0.00). Estimates in shores were smaller and inconsistent.

### Differentially methylated CpG sites

At FDR < 0.05, we identified 469 CpGs for CRII, 752 for neutrons, 6,392 for SSN, and 46 for IMF out of 360,272 tested sites (**Table S1-4**). Many of the strongest signals mapped to genes with known roles in genome maintenance and aging biology, including *ZNF365* (DNA damage response and replication stress), *BCL2L11/BIM* (apoptosis), *GRB10* (insulin/IGF and growth-factor signaling), and *PANX2* (neuronal and axon biology). The Kp-index yielded no significant CpGs and being null at summary level associations, was thus excluded from downstream analyses.

Of SSN-associated CpGs, 5,727 (87.6%) were unique; CRII had 27 (5.8%) unique, neutrons 65 (8.6%), and IMF 2 (4.3%), indicating partly distinct epigenetic fingerprints (**Fig. 1c**). Furthermore, the direction of methylation change followed clear exposure-specific patterns: CRII and neutrons were predominantly associated with lower methylation (86 and 87%, respectively), whereas SSN and IMF were predominantly associated with higher methylation (93 and 94%, respectively) (**Fig. 1d**). Notably, CpG sites showing higher methylation with SSN overlapped substantially with those showing lower methylation with CRII and neutrons (**Fig. S5a**). Significant CpGs were enriched in CpG islands (gene regulatory regions) and OpenSea regions (isolated sites in the genome) (**Fig. S5b**).

### Differentially methylated regions (DMR)

DMRs represent clusters of neighboring CpG sites that show coordinated methylation differences, thus can be more biologically robust than single-site changes. Using comb-p (seed p = 0.001; max gap = 1 kb; ≥ 3 CpGs; region FDR < 0.05; hg19) applied to CpGs reaching FDR < 0.05 in single-site analyses, we identified 704 genomic regions that were differentially methylated for SSN, 4 for IMF, 42 for CRII, and 67 for neutrons (**Fig. 2a-d; Table S5-8**), with strong island enrichment (**Fig. S5c**). Several loci showed consistent signals across multiple exposures, and 33 regions were shared by SSN, CRII, and neutrons (**Fig. 3a**).

**Figure 2.**
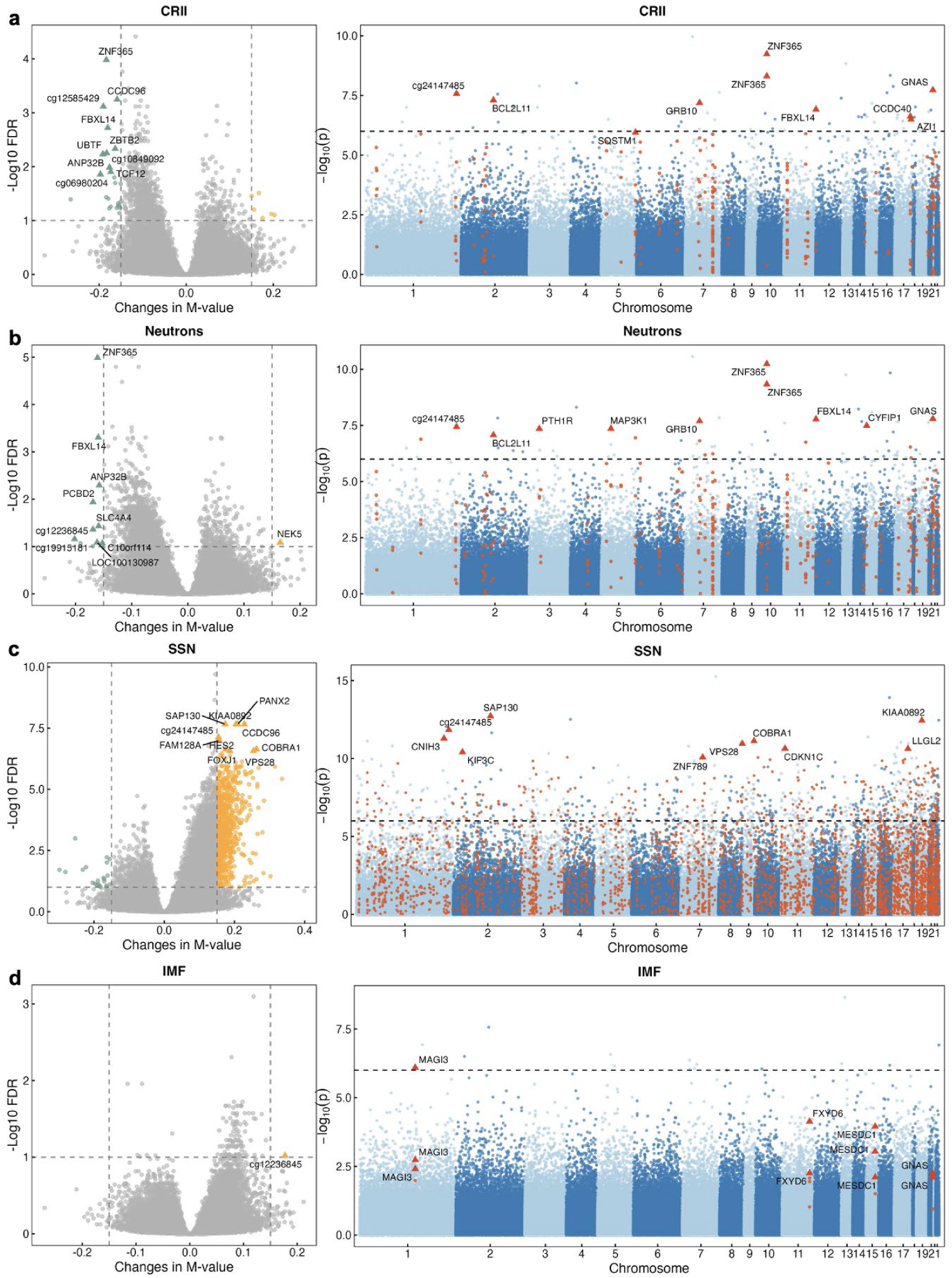
CpG-level and regional DNA methylation signals associated with space-weather exposures. Left panels: Volcano plots showing associations between space-weather indicators and methylation at individual CpG sites (differentially methylated positions, DMPs). The x-axis represents estimated changes in logit-transformed methylation values (M-values), and the y-axis represents –log10 false discovery rate (FDR). Sites with absolute methylation change >0.15 and FDR <0.1 are highlighted, with higher-methylation sites in orange and lower-methylation sites in green. **Right panels**: Differentially methylated regions (DMRs) identified using comb-p (seed *p* = 0.001, maximum distance = 1 kb, ≥ 3 CpGs, region-level FDR < 0.05), with per-CpG association statistics shown to illustrate region-level concordance. All analyses used IQR-scaled 30-day moving averages of exposures. Models were adjusted for age, year, blood-cell composition, batch factors, smoking status, pack-years, body mass index, education, diabetes, antihypertensive medication, statin use, temperature, visibility, and season.

**Figure 3.**
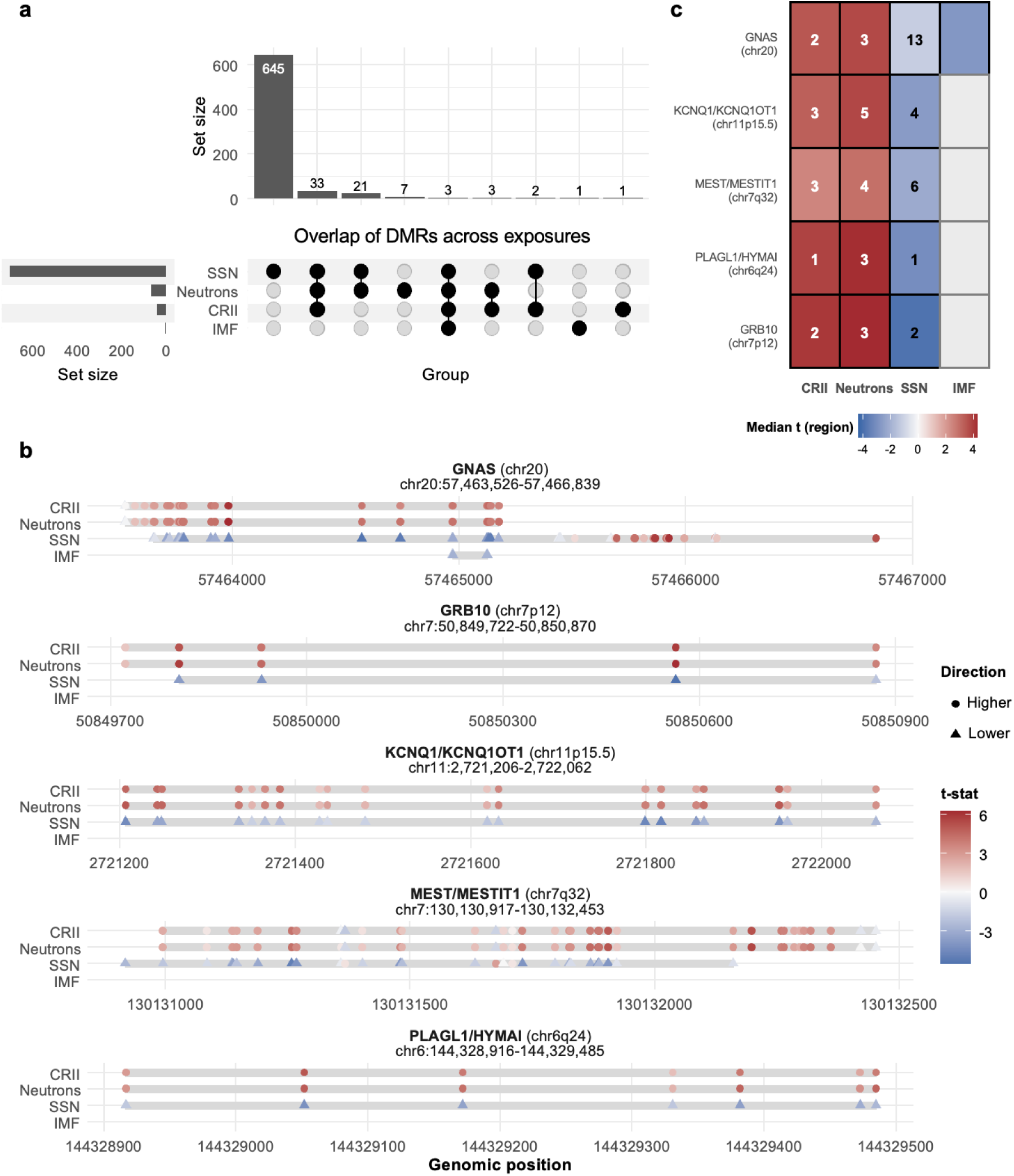
DNA methylation patterns at canonical imprinted domains across space-weather exposures. (**a**) Overlap of significant DMRs across exposures, shown by UpSet plot. DMRs were identified using comb-p (seed *p* = 0.001, maximum distance = 1 kb, ≥ 3 CpGs, region-level FDR < 0.05). (**b**) Locus-specific tracks at five canonical imprinted domains (*GNAS* [20q13.32], *KCNQ1/KCNQ1OT1* [11p15.5], *PLAGL1/HYMAI* [6q24], *MEST/MESTIT1* [7q32], and *GRB10* [7p12]). Panels show genomic coordinates (hg19), CpG-level test statistics (t values) for each exposure, and direction of methylation change (higher methylation = circles, lower methylation = triangles). (**c**) Concordance across exposures at imprinted domains. Numbers indicate the count of significant differentially methylated regions (DMRs) within each locus; color shading reflects the median t-statistic across CpGs.

Among these, several top DMRs were localized within canonical imprinted domains, genomic sites where DNAm normally regulates expression of only one parental copy of a gene and is usually kept stable.^40^ These included *GNAS* (20q13.32; 26-42 CpGs; detected with all four indicators), *KCNQ1/KCNQ1OT1* (11p15.5; 19 CpGs; detected with CRII, neutrons, SSN), *PLAGL1/HYMAI* (6q24; 7 CpGs; detected with CRII, neutrons, SSN), *MEST/MESTIT1* (7q32; 28-37 CpGs; detected with CRII, neutrons, SSN), and GRB10 (7p12; 4-5 CpGs; detected with CRII, neutrons, SSN) (**Fig. 3b**).

At these domains, the changes were modest but consistent in direction: CpGs tended to show higher methylation with GCR indicators and lower methylation with SGA indicators. This pattern contrasted with the genome-wide trend (GCRs overall associated with lower methylation and SGAs with higher methylation), warranting further investigation (**Fig. 3c**).

### Pathway enrichment

Bias-aware methylGSA identified both shared and exposure-specific enriched biological pathways (**Fig. 4, 5**). Across indicators, we consistently saw signals related to DNA replication and repair, cell-cycle control, circadian regulation, and energy metabolism. These shared pathways suggest a broad impact on genome maintenance and cellular stress responses. Against this shared background, CRII and neutrons showed stronger enrichment of DNA damage and repair processes, together with inflammatory responses. In contrast, SSN and IMF showed stronger enrichment for protein handling and metabolic control, including unfolded protein response, PI3K-AKT-mTOR signaling, and protein translation.

**Figure 4.**
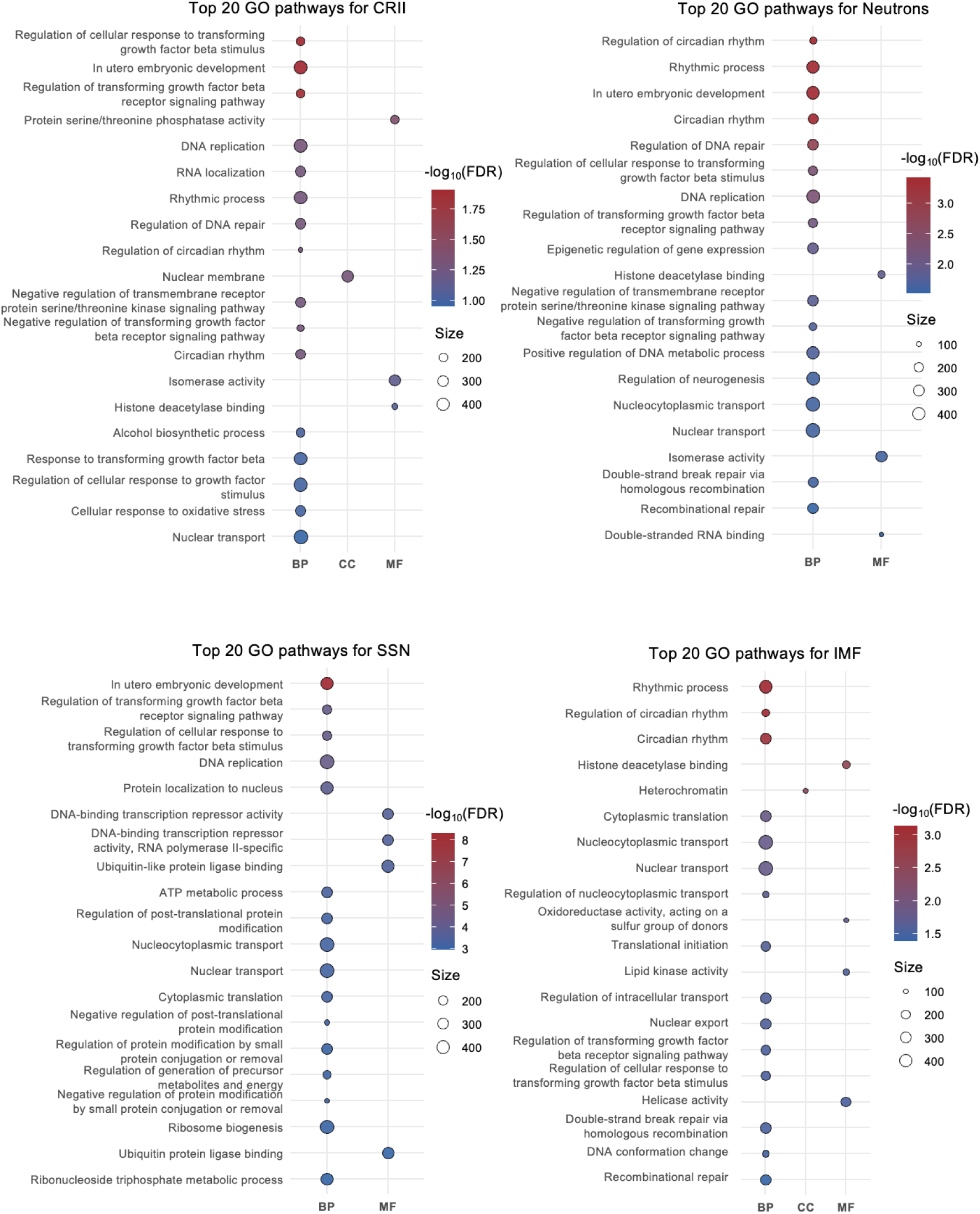
Gene Ontology (GO) enrichment analysis of CpG-level EWAS results. Top 20 enriched GO terms are shown for CRII, neutrons, SSN, and IMF. Terms are grouped by GO domain: Biological Process (BP), Cellular Component (CC), and Molecular Function (MF). Point size indicates the number of genes in each set, and color intensity reflects statistical significance (–log10 FDR). Enrichment analyses were performed using methylGSA, with CpG-level FDR values from EWAS as input and Benjamini-Hochberg correction applied across pathways.

**Figure 5.**
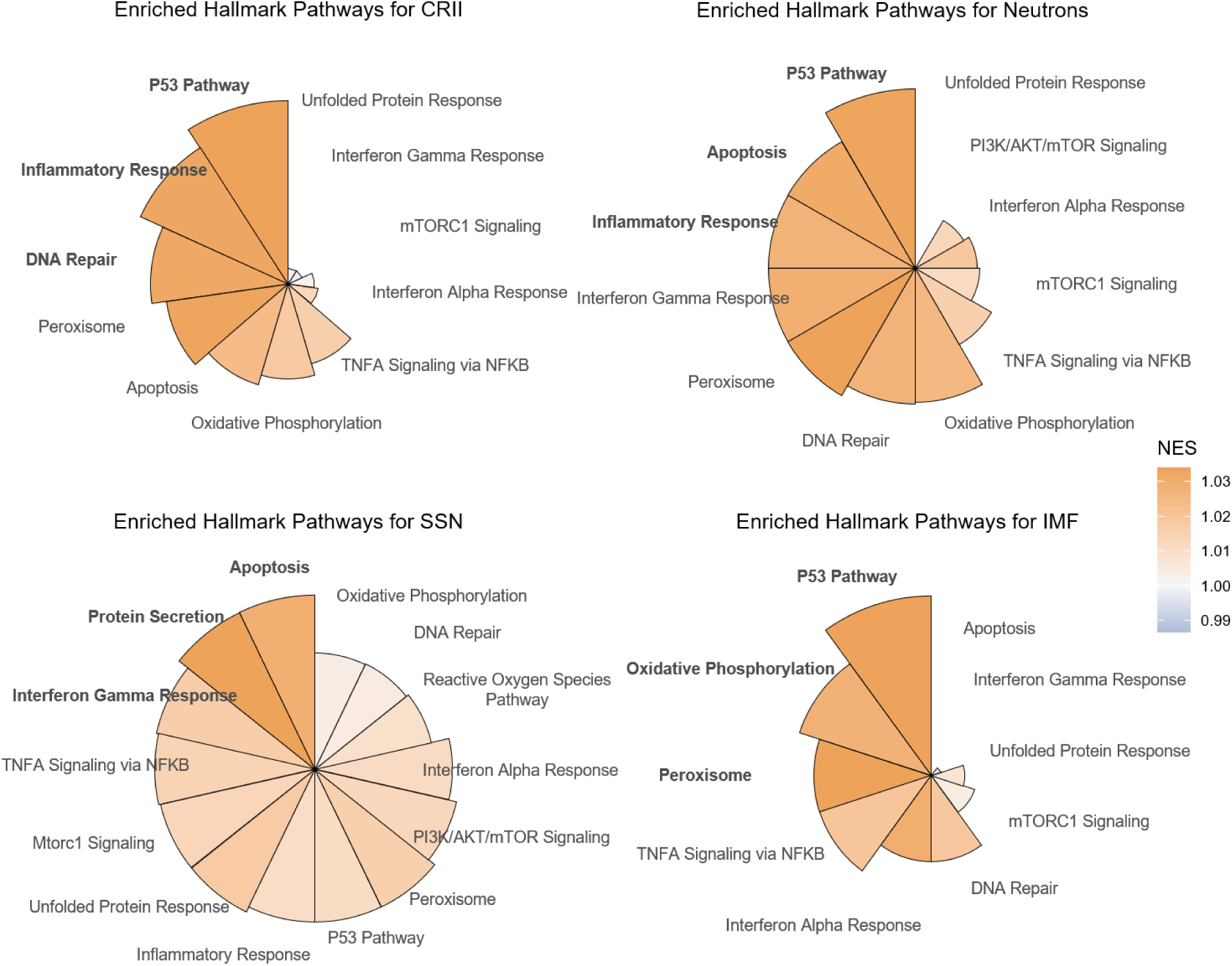
Hallmark pathway enrichment and associations between space-weather exposures and epigenetic age acceleration. Enrichment of MSigDB Hallmark gene sets based on CpG-level EWAS results. Normalized enrichment scores (NES) are shown for the top pathways across CRII, neutrons, SSN, and IMF.

Because EWAS effects were directionally asymmetric, we also ran direction-split analyses separating CpGs associated with higher versus lower methylation. Splitting reduced power but unmasked sign-specific biology indicating that neurodevelopment and axon guidance pathways became prominent, especially for SSN-associated higher methylation, and for neutrons in both higher and lower methylation subsets (**Fig. S6**). In Hallmarks, GCRs (CRII and neutrons) linked lower methylation showed apoptosis enrichment, whereas CRII-associated higher methylation highlighted PI3K-AKT-mTOR signaling. SGAs (SSN and IMF) associated lower methylation highlighted protein secretion and IMF-associated higher methylation showed P53 and DNA repair involvements (**Fig S7**). Normalized enrichment scores (NES) were small (1.0–1.03), reflecting subtle but coordinated shifts across thousands of CpGs rather than strong single-pathway effects.

## Discussion

In a longitudinal cohort of elderly and mostly white men, short-term variability in space weather was associated with distinct epigenomic changes. GCRs (CRII and neutrons) were associated with higher EAA, while SGAs (SSN and IMF) were associated with lower EAA. Furthermore, GCRs were associated with altered DNAm in pathways related to DNA damage response, p53 signaling, and inflammation, whereas SGAs were linked to altered DNAm in secretory and metabolic pathways. These findings together extend population-based evidence that motivated this study, showing that natural fluctuations in the space environment leave measurable epigenomic signatures of genome integrity and aging in blood, detectable within 30-day exposure windows. In line with previous findings in astronauts and space crews showing biological aging and methylation changes during missions,^16,41^ our results suggest that ground population might also be biologically responsive to space weather fluctuations and less protected from their influences than previously assumed.

Our analysis with DNAm clocks extends our earlier telomere studies by showing that GCR exposures, previously linked to shorter telomeres, are associated with accelerated epigenetic aging, while SGAs, previously linked to longer telomeres, are associated with slower aging.^7^ The EAA metrics derived from Horvath, Hannum, and PhenoAge clocks all moved in the same directions: higher with GCRs and lower with SGAs. The Horvath clock and Hannum clock are DNAm predictors of chronological age,^9,42^ while the PhenoAge clock is derived from clinical biomarkers and is more closely related to inflammatory and physiologic dysregulation.^24^ By contrast, GrimAge is trained to predict mortality based on smoking- and protein-related surrogates.^25,42^ GrimAgeAccel showed no clear association here, and is plausibly less sensitive to short 30-day variability in this cohort, whereas the other clocks captured short-term patterns. These findings suggest epigenetic clocks might serve alongside telomere length as complementary biomarkers of radiation-linked biological aging and are more sensitive to short-term changes.

The clock patterns were mirrored in EWAS signals and global DNAm metrics. GCRs were associated with lower methylation at the majority (86−87%) of CpGs reaching statistical significance and with reduced methylation entropy in CpG islands; whereas SGAs exhibited higher methylation at most (93−94%) significant CpGs accompanied by increased entropy. Previous studies have reported GCR components such as ^28^Si and X-ray induce decreases in DNAm, supporting our findings in the same direction.^43^ These coherent yet small methylation shifts across many loci are consistent with previous observations that global DNAm changes during spaceflight are modest and often reversible. For example, in the NASA Twins Study, most DNAm changes observed during the mission were smaller than interindividual variability and largely returned to baseline post flight.^16^ Reduced island entropy under GCR implies more constrained promoter states under ionizing radiation stress, consistent with enrichment of DNA repair, P53, and apoptosis pathways.^44^ This aligns with previous reports of GCR-induced DNA damage responses and selective hypomethylation in radiation-sensitive genomic contexts.^45^ In contrast, higher island entropy under SGA suggest heterogeneous promoter methylation. The null findings for Kp index indicate that higher-energy electromagnetic radiation related to solar activity indices (SSN and IMF) and GCR proxies can potentially promote variations in blood DNAm. It is important to note, however, that our results are based on bulk DNAm, lacking cell-type specific resolution, which astronaut multi-omic studies suggest may capture heterogeneous and lineage-specific responses.^14,16^

Several of the top genomic regions that were differentially methylated across the space weather indicators localized within or near well-established human imprinted gene domains. These domains are governed by parent-of-origin regulation, with regions where methylation is set differently on maternal versus paternal copies, often influencing the activity of multiple nearby genes. Interestingly, the direction of methylation changes at these regions often flipped compared with the broader genome-wide patterns, suggesting that imprinted domains may be regulated differently from the rest of the genome. However, given the limited statistical power, these findings should be considered exploratory and warrant replication in cell-sorted or single-cell methylome studies. Many of the affected domains harbor genes central to growth and metabolism (e.g., *GNAS* in G-protein hormone signaling,^46^ *GRB10* in growth control and insulin signaling;^47^ and *KCNQ1* in development and heart function^48^). While our data cannot establish parent-of-origin changes, the repeated involvement of these loci suggests imprinted domains as sensitive genomic regions worth further study.

Pathway analyses revealed that the space weather indicators were associated with physiological stress responses involving immune, repair, and metabolic pathways, but along with different trajectories. GCR exposures preferentially showed enrichment in DNA damage response, p53 signaling, and inflammatory pathways, a pattern that mirrors findings from both the NASA Twins Study and SOMA multi-omics, which reported radiation-associated DNA damage responses and immune activation.^14,16,45^ These results are further supported by experimental evidence showing that ionizing radiation induces methylation changes enriched in DNA repair and apoptosis pathways.^44^ In contrast, SGA exposures more strongly implicated metabolic and protein-handling process, including oxidative phosphorylation, unfolded-protein response, and protein secretion, consistent with metabolic and secretory adaptations reported across SOMA investigations. Given that enrichment analysis reflects over-representation rather than direct activation, and the NES values were modest, these signatures likely represent coordinated, modest shifts across many loci rather than large changes in a single pathway, yet they are directionally consistent with the global methylation patterns and with the contrasting aging trajectories observed for GCRs versus SGAs.

Beyond genome integrity and metabolism, additional enrichment signals pointed to brain- and circadian-related processes. Neurodevelopment and neurogenesis pathways appeared only when CpG sites with higher and lower methylation were analyzed separately. Although exploratory, these direction-specific findings might be especially relevant in our elderly cohort, where participants are more vulnerable to cognitive decline and neurodegenerative risk. This may help explain prior findings in the same cohort showing that higher SSN was linked to poorer cognitive function.^2^ In addition, SOMA reported brain-associated proteins in plasma and broad circadian and sleep-related perturbations across missions, providing external biological plausibility.^45^ Furthermore, circadian rhythmic pathways were most enriched under GCRs and IMF, broadly aligning with previous studies linking SGAs and GCRs to circadian rhythms and sleep physiology.^6,49,50^ The findings together imply the possibility that space-weather variability influence neuronal and circadian biology, although further mechanistic explanations are needed.

We acknowledge several limitations of our study. First, we studied methylation in blood, which may not fully reflect changes in other tissues. Nonetheless, blood is widely used as a surrogate in population studies and most epigenetic clocks have been developed from and mostly used on blood DNA; also, epigenetic clocks such as Horvath’s are pan-tissue measures of systemic aging. Integration of other omics layers, particularly proteomics, could strengthen functional interpretation. Second, residual time-varying confounding (e.g., co-occurring environmental exposures) cannot be fully excluded, although the consistency of findings across multiple covariate-adjusted models suggests this bias is unlikely to be large. Third, our participants were all older men from a region of relatively high socioeconomic status. While this limits generalizability, older adults are already vulnerable to environmental stressors and accelerated aging, making them an important population to study. Further studies should extend to women, other ethnic and racial groups, younger ages, and other sensitive populations such as pregnant individuals or people with chronic conditions. Fourth, our space-weather exposures were global indices measured over the same time windows, making it difficult to disentangle the independent effects of SGAs and GCRs, which are inherently anticorrelated.^39^ Experimental or longitudinal studies spanning multiple solar cycles will be needed to clarify their independent causal contributions. Moreover, these global indices do not capture personal-level dose. Stronger associations may be detectable in high-altitude populations, or with finer geomagnetic indices and personal-level proxies.

Our study has notable strengths. Our repeated-measures design, spanning more than one full 11-year solar cycle, allowed us to assess space weather fluctuations in a longitudinal framework. By integrating epigenetic clocks, EWAS, regional analyses, and pathway enrichment, we provide a comprehensive picture of how short-term space weather variability influences biological aging signatures in the epigenome and uncovered biologically meaningful pathways that may help explain phenotype-level associations observed in populations. Detailed participant information and environmental information enabled rigorous covariate adjustment, and bias-aware statistical approaches minimized inflation and false discovery. Taken together, these features increase confidence in the robustness of our results.

Our findings suggest that even at ground level, space weather variability may influence biological aging and contribute to population health risks. From a public health perspective, this highlights space weather as a largely overlooked environmental factor that may interact with aging trajectories and chronic disease risk. Replication in independent and more diverse cohorts, along with functional follow-up using multi-omics, will be essential to validate and extend these observations.

## Data Availability Statement

DNA methylation data analyzed in this study are available through controlled access at dbGaP under accession number phs000853.v2.p2. Analysis code used in this study is hosted at https://github.com/zhaiting/NAS_DNAm and will be made publicly available upon publication. All other data supporting the findings of this study are available from the corresponding author upon reasonable request.

## Acknowledgments

The authors would like to thank the NAS study participants for their dedicated participation.

## Ethical Approval

The VA normative aging study was approved by the institutional review boards of the Harvard T.H. School of Public Health and the Veterans Administration Boston Healthcare System, and all participants provided written informed consent.

## Author contributions

Conceptualization: TZ, CLZV, JS, PK

Methodology: TZ, CLZV

Investigation, Formal analysis, Visualization, Writing – original draft: TZ

Funding acquisition: JS, AAB, PV, ZDN, PK

Resources: CLZV, PV, JS, PK

Writing – review & editing: all authors

## Supporting Information for

**Figure S1.**
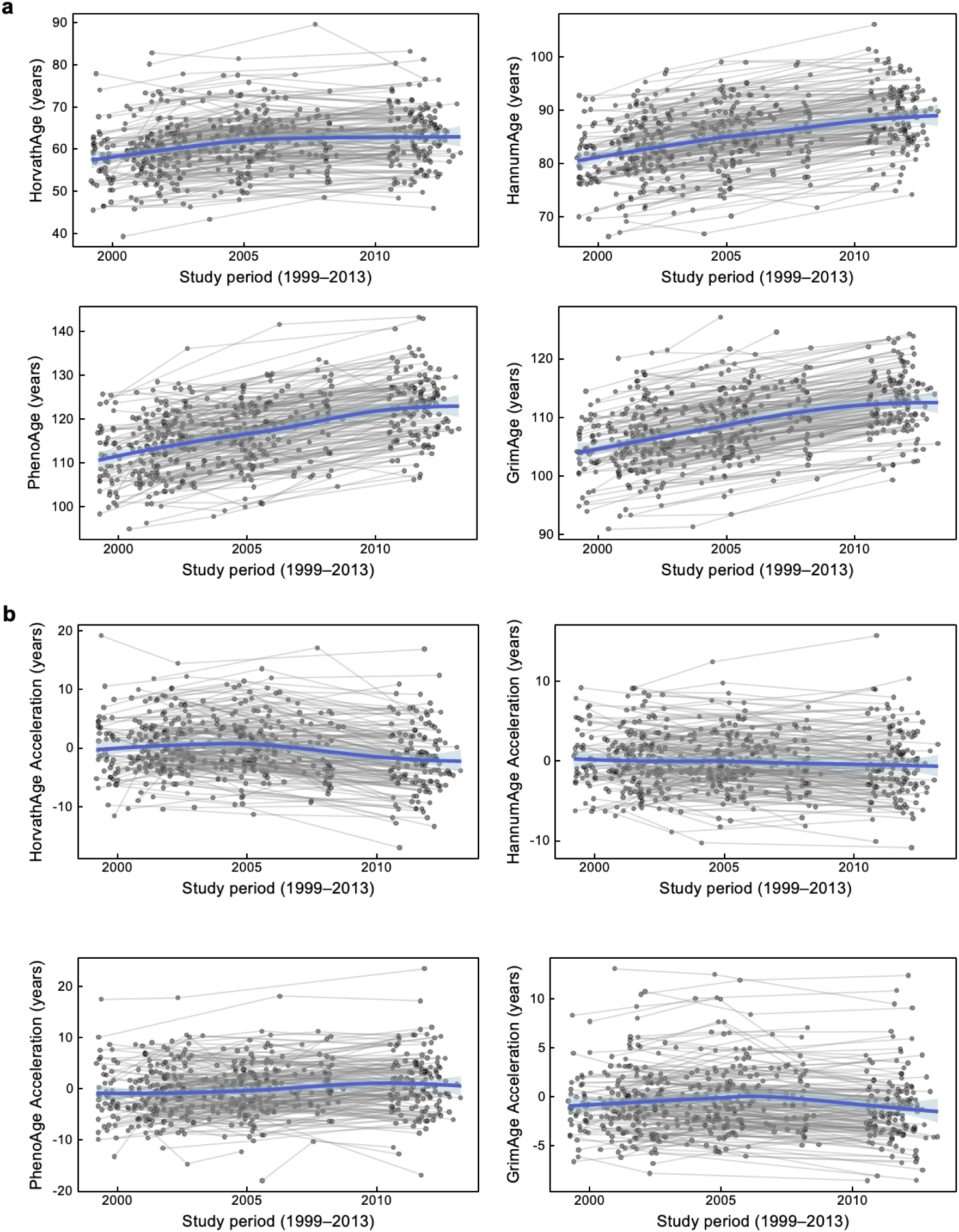
Temporal patterns of epigenetic aging measures. (**a**) Individual trajectories of epigenetic aging clocks (HorvathAge, HannumAge, PhenoAge, GrimAge) and (**b**) corresponding acceleration metrics among participants with at least three repeated samples (1999–2013). Lines connect samples from the same individual, and blue lines indicate LOESS fits with 95% confidence intervals.

**Figure S2.**
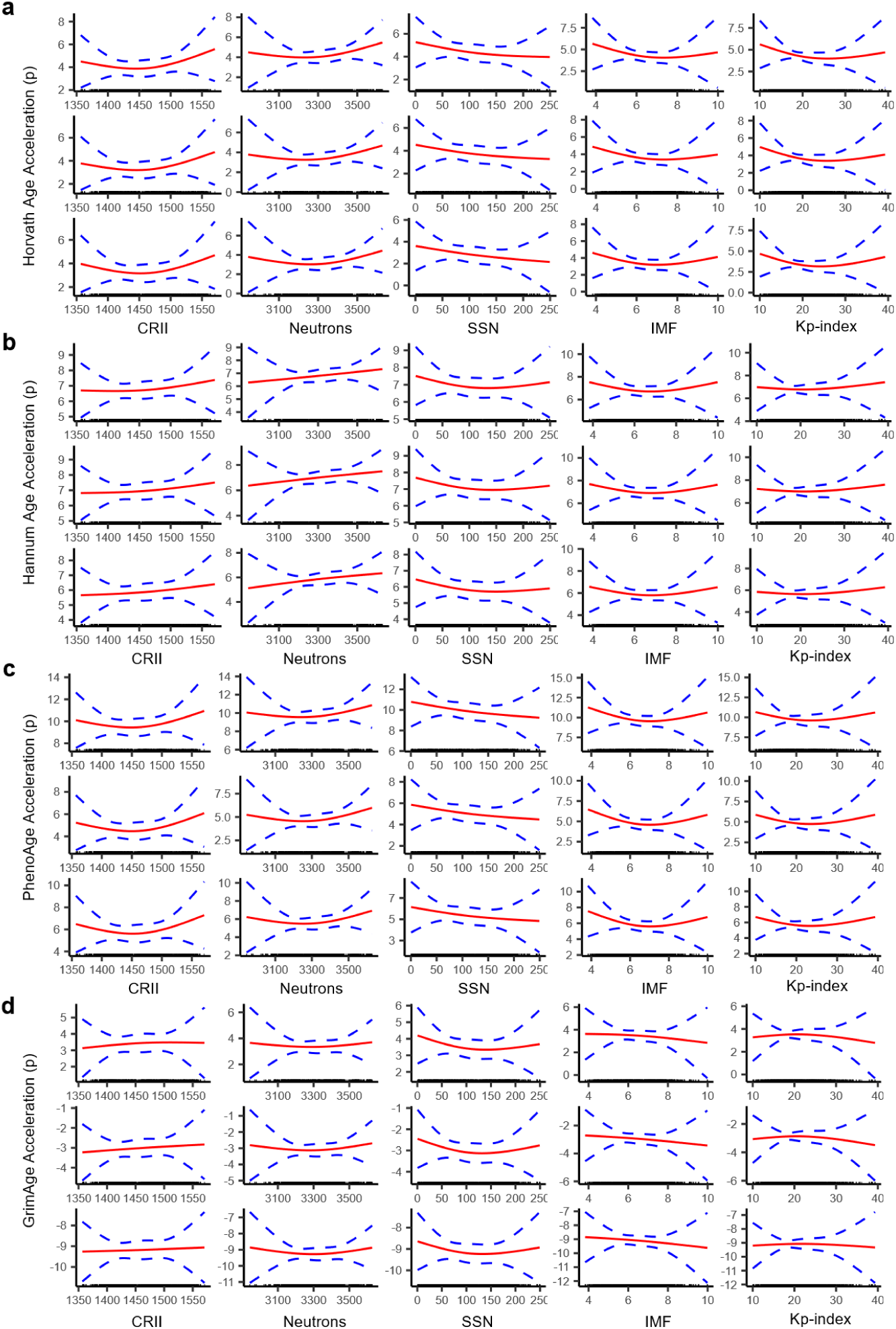
Non-linear exposure-response associations between space-weather exposures and epigenetic age acceleration. For each exposure, CRII (cosmic ray-induced ionization), neutrons, SSN (sunspot number), IMF (interplanetary magnetic field), and Kp-index, associations with acceleration of four epigenetic clocks are shown: (**a**) HorvathAge, (**b**) HannumAge, (**c**) PhenoAge, and (**d**) GrimAge. Curves were modeled using natural cubic splines with 3 degrees of freedom. Solid red lines represent predicted associations, and dashed blue lines denote 95% confidence intervals (CIs). Within each panel, models were specified sequentially: top subpanels adjusted for age, year, blood-cell composition, and batch factors; middle subpanels additionally adjusted for smoking status, pack-years, body mass index (BMI), education, diabetes, antihypertensive medication, and statin use; bottom subpanels further adjusted for temperature, visibility, and season. Exposures were scaled per interquartile range (IQR) of their 30-day moving averages.

**Figure S3.**
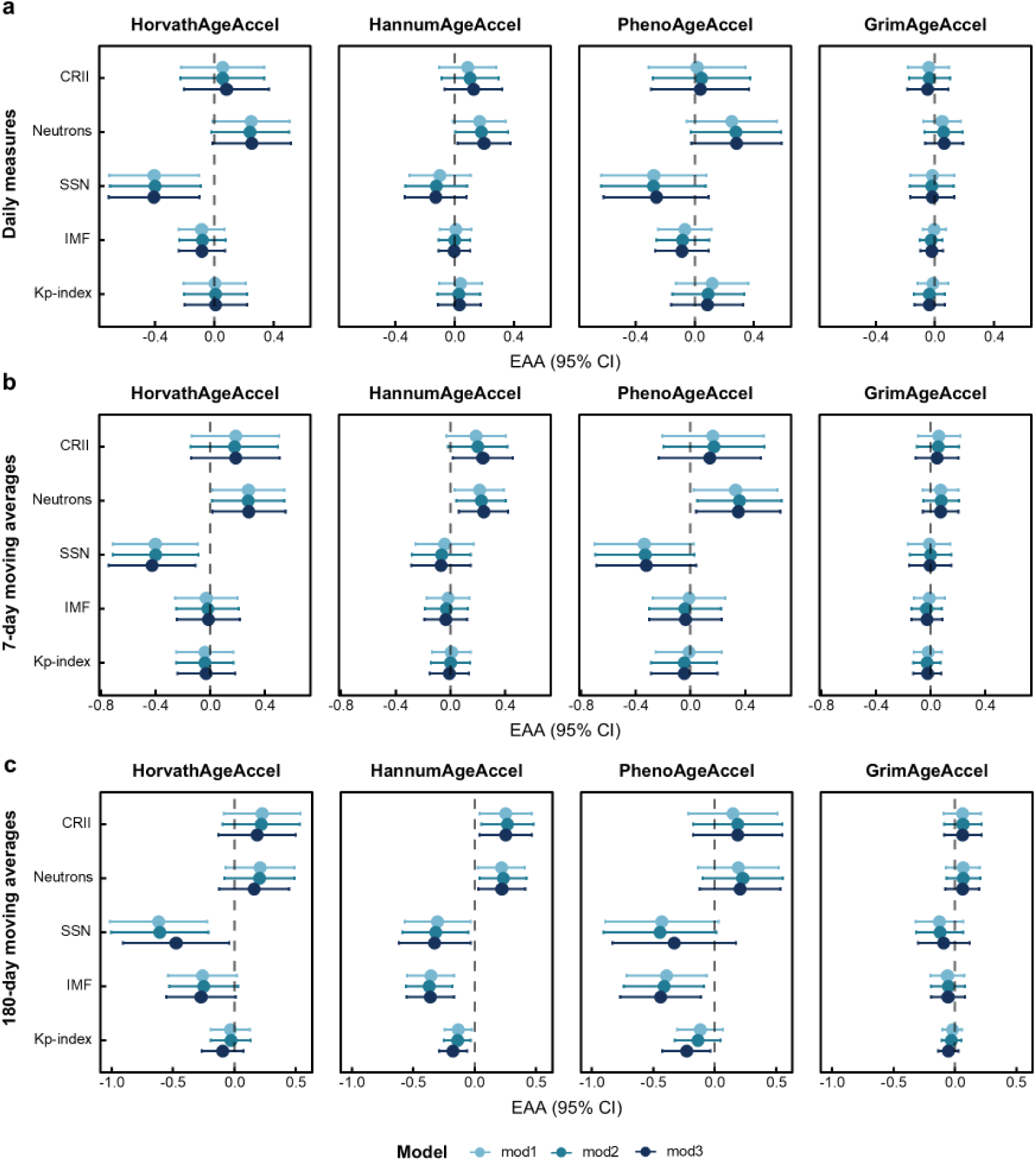
Sensitivity analyses of associations between space-weather exposures and epigenetic age acceleration (EAA). Forest plots show associations (β per interquartile range [IQR] with 95% confidence intervals [CIs]) between exposures, CRII (cosmic ray-induced ionization), neutrons, SSN (sunspot number), IMF (interplanetary magnetic field), Kp-index, and EAA metrics from four clocks: HorvathAgeAccel, HannumAgeAccel, PhenoAgeAccel, and GrimAgeAccel. Results are presented for three exposure averaging windows: (**a**) daily values, (**b**) 7-day moving averages, and (**c**) 180-day moving averages. Models were specified sequentially: mod1 adjusted for age, year, blood-cell composition, and batch factors; mod2 additionally adjusted for smoking status, pack-years, body mass index (BMI), education, diabetes, antihypertensive medication, and statin use; mod3 further adjusted for temperature, visibility, and season. Exposures were scaled per IQR.

**Figure S4.**
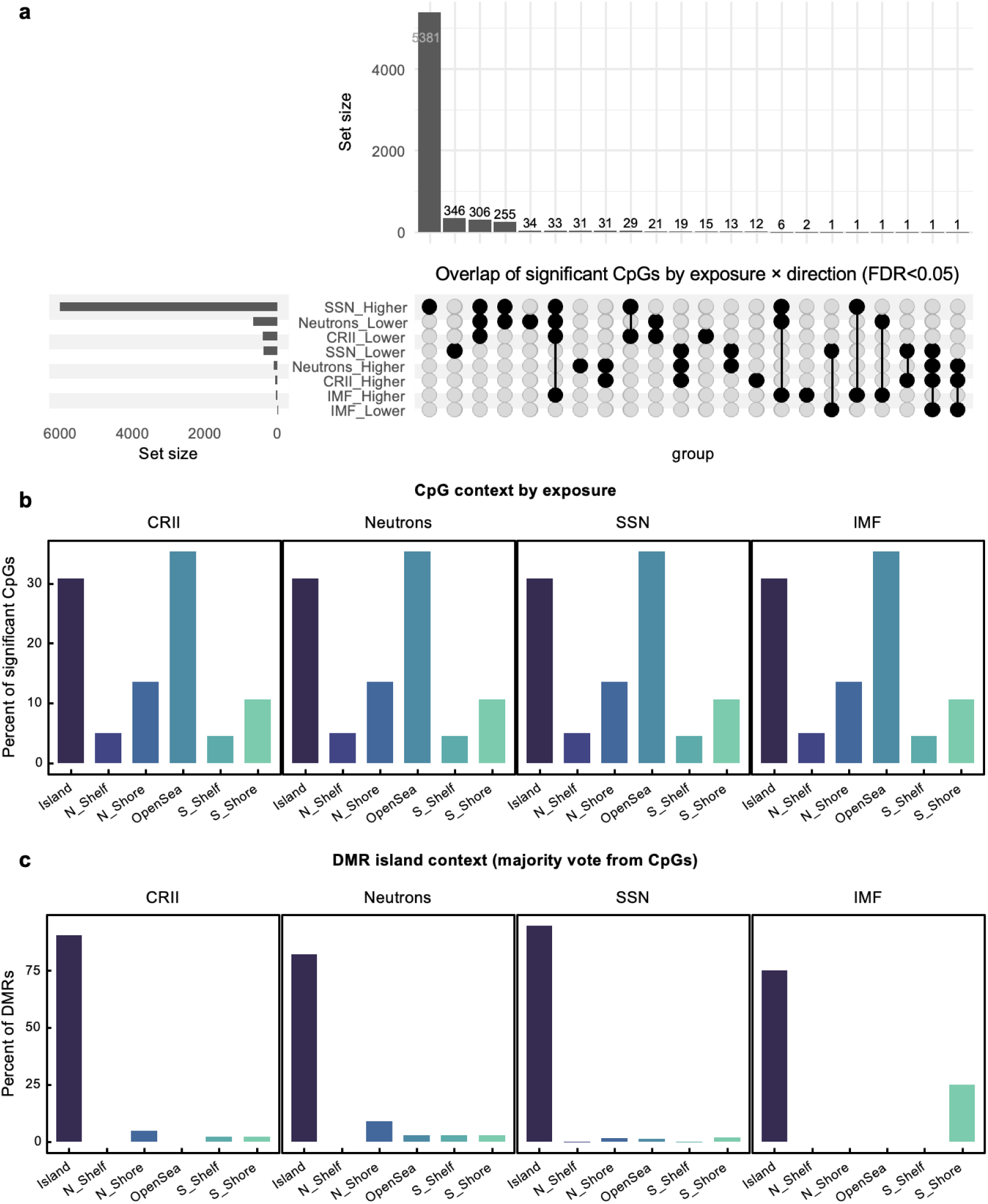
Structure of EWAS signals across space-weather exposures. **(a)** Overlap of significant CpGs (false discovery rate <0.05) by exposure and methylation direction (lower vs higher methylation). **(b)** Distribution of significant CpGs by CpG-island annotations (islands, shores, shelves, OpenSea). **(c)** Proportion of differentially methylated regions (DMRs) in which the majority of contributing CpGs were located in CpG islands.

**Figure S5.**
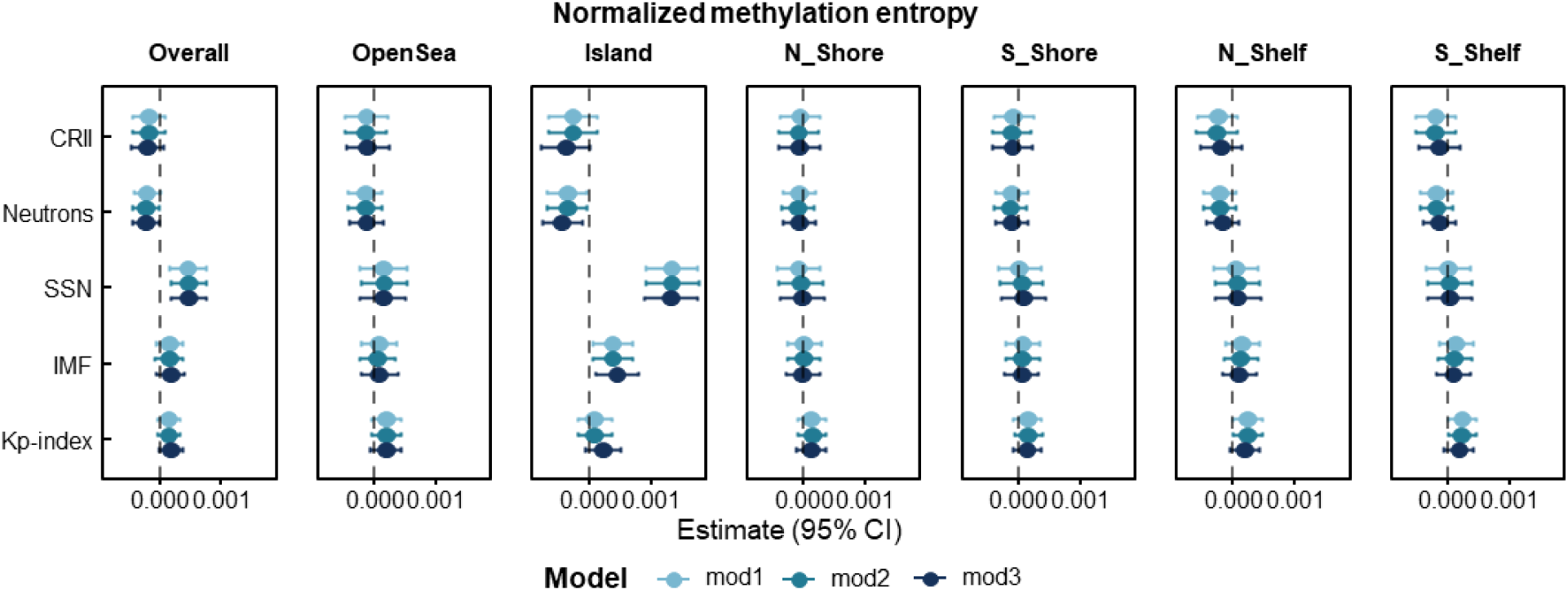
Associations between space-weather exposures and methylation entropy. Forest plots show associations (β per interquartile range [IQR] with 95% confidence intervals [CIs]) between exposures and normalized methylation entropy, overall and stratified by genomic context (OpenSea, island, north/south shore, north/south shelf). Models were specified sequentially: mod1 adjusted for age, year, blood-cell composition, and batch factors; mod2 additionally adjusted for smoking status, pack-years, body mass index, education, diabetes, antihypertensive medication, and statin use; mod3 further adjusted for temperature, visibility, and season. Exposures were scaled per interquartile range (IQR) of 30-day moving averages.

**Figure S6.**
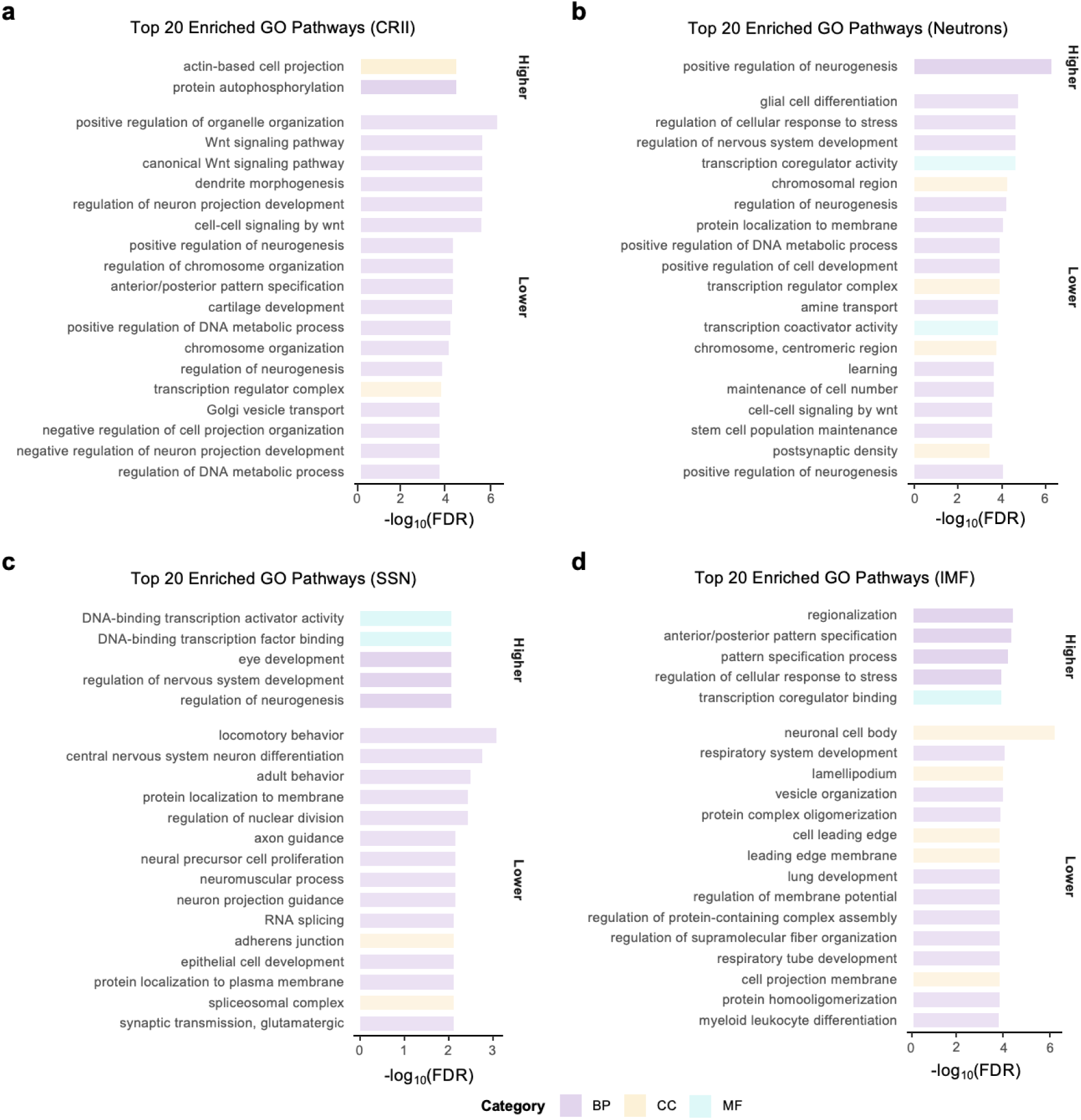
Direction-stratified Gene Ontology (GO) enrichment analysis across space-weather exposures. For each exposure, (**a**) CRII (cosmic ray-induced ionization), (**b**) neutrons, (**c**) SSN (sunspot number), and (**d**) IMF (interplanetary magnetic field), the top 20 enriched GO terms are displayed separately for CpGs associated with higher or lower methylation (false discovery rate [FDR] < 0.05). Bars represent enrichment significance as –log_10_ (FDR). GO terms are grouped by domain: Biological Process (BP), Cellular Component (CC), and Molecular Function (MF).

**Figure S7.**
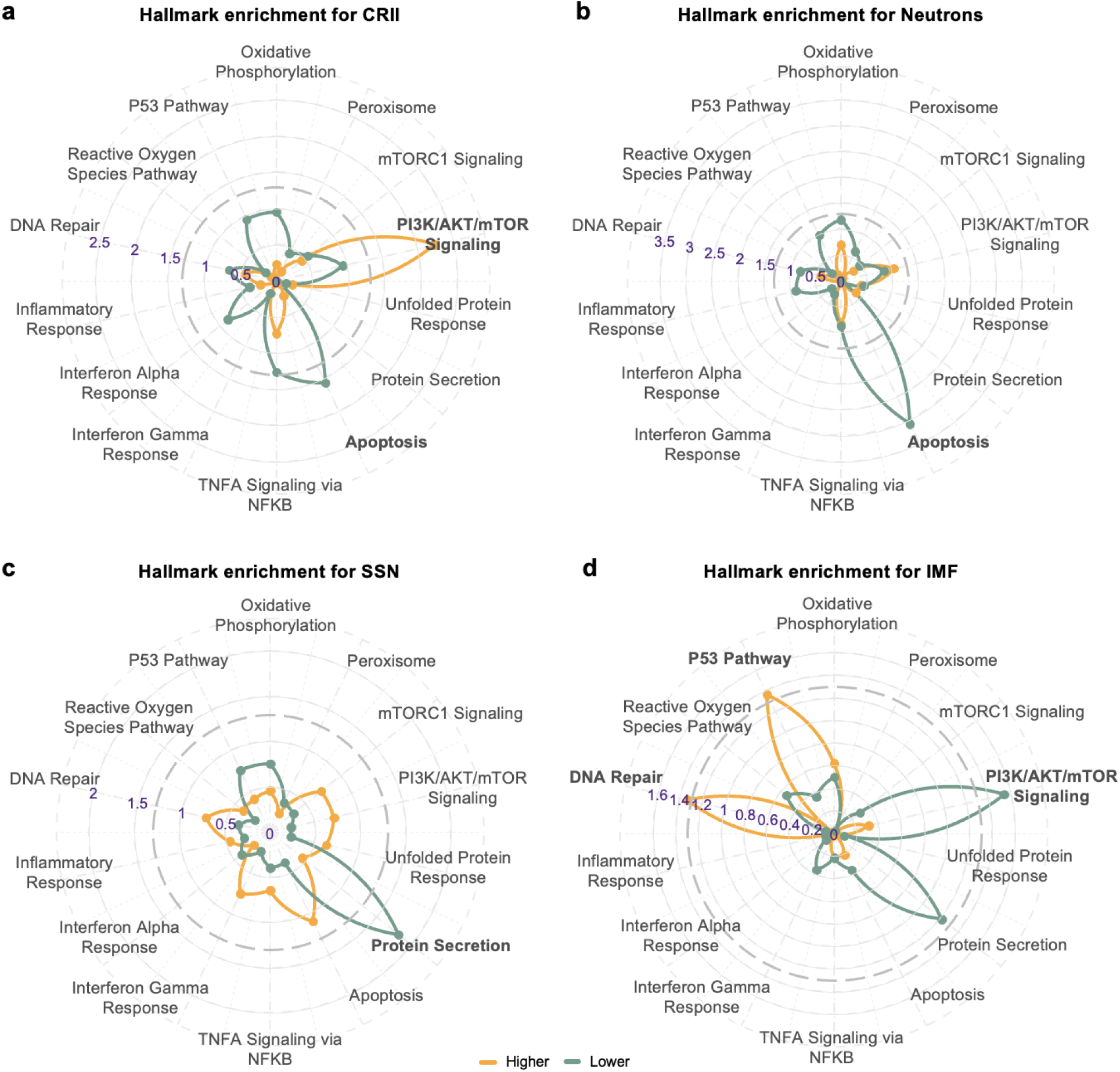
Direction-stratified Hallmark enrichment analysis across space-weather exposures. For each exposure, (**a**) CRII (cosmic ray-induced ionization), (**b**) neutrons, (**c**) SSN (sunspot number), and (**d**) IMF (interplanetary magnetic field), Hallmark gene set enrichment results are shown separately for CpGs associated with higher methylation (orange) or lower methylation (green). Axes represent normalized enrichment scores (NES), with pathways arranged radially.

